# The national, longitudinal NASCITA birth cohort study: prevalence of overweight or obesity at 12 months of age in children born healthy

**DOI:** 10.1101/2022.07.14.22277595

**Authors:** Antonio Clavenna, Eleonora Morabito, Massimo Cartabia, Rita Campi, Chiara Pandolfini, Maurizio Bonati, the NASCITA Working Group

## Abstract

**Objective:** To estimate the prevalence of overweight/obesity at 12 months in an Italian birth cohort and to identify factors related to an increased likelihood of being overweight.

**Methods:** The Italian NASCITA birth cohort was analysed. Infants were classified as underweight (<5^th^), normal weight (5^th^-85^th^), and overweight (>85^th^ centile) at 12 months of age according to the World Health Organization percentiles of body mass index and the prevalence of overweight/obesity was estimated. To test the association between the chance of being overweight/obese and parental and newborn characteristics, and infant feeding, healthy newborns (no preterm/low birth weight, and with no malformations), with appropriate-for-gestational-age birth weight were selected, and univariate and multivariate analyses were performed

**Results:** The prevalence of overweight was 23.5% (95%CI 22.2-24.8) in the 4,270 neonates followed during their first year of life, and 23.7% (631/2749) in healthy newborns with a birth weight that was appropriate for gestational age.

A big infant appetite (Odds Ratio 4.39, 95%CI: 2.73-7.07), living in southern Italy (OR 1.64, 95%CI: 1.35-2.00) and maternal age <30 years (OR 1.39, 95%CI: 1.05-1.83) were the main variables associated with a greater likelihood of being overweight/obese. Exclusive breastfeeding for at least 6 months did not influence the chance of being overweight.

**Conclusions:** The sociodemographic factors (e.g. area of residence, age) seem to be the most relevant determinants influencing the chance of being overweight. In order to prevent childhood and adult obesity, early interventions should be set up or improved, with particular attention to vulnerable families.

## INTRODUCTION

Childhood overweight and obesity represent an increasing public health issue that threatens future health and quality of life.[1] Evidence suggests that there are early-life factors (e.g. maternal body mass index, gestational weight gain, maternal smoking habits, and infant feeding) associated with the development of obesity.[2,3] Nutrition, in particular, plays an important role in growth and development, and breastfeeding is considered by the World Health Organization (WHO) the best method of nourishment for infants for its positive health effects on children and mothers in the short and long term.[4]

For these reasons the WHO recommends that children should be exclusively breastfed for the first 6 months of life; afterward, they should begin eating safe and adequate complementary foods while continuing breastfeeding for up to 2 years and beyond.[4]

Several studies found an association between breastfeeding and a reduced risk of being overweight, even if results are not conclusive.[5–7]

Italy is one of the European countries with the greatest prevalence of childhood overweight and obesity, with a rate in 8-year-old children of 42% and 21%, respectively.[8,9] A North-South gradient was detected not only in Europe, but also within Italy.[10] In particular, school aged children living in the South of Italy have a 2-fold greater risk of overweight compared to children living in the North. [10]

However, there is scant information concerning the proportion of overweight/obese Italian children at an early age.

In this context, this study aimed to describe the growth of newborns participating in the Italian national NASCITA cohort in their first year of life, to estimate the prevalence of overweight/obesity at 12 months of age, and to identify potential factors associated with a greater likelihood of being overweight.

## METHODS

### Data source

The NASCITA birth cohort was set up by the Laboratory for Mother and Child Health of the Istituto di Ricerche Farmacologiche Mario Negri IRCCS in Milan in collaboration with the national Paediatric Cultural Association (ACP).

The methods of the NASCITA cohort have been described elsewhere.[11,12] Briefly, Italian health care is provided free or at a minimal charge. All Italian children receive primary health care exclusively from a family paediatrician until they are at least six years old as part of universalistic health system organization. Seven well-child visits are scheduled by the paediatrician in the first six years of a child’s life to monitor growth and development and offer preventive care (Figure 1).

**Figure 1.**
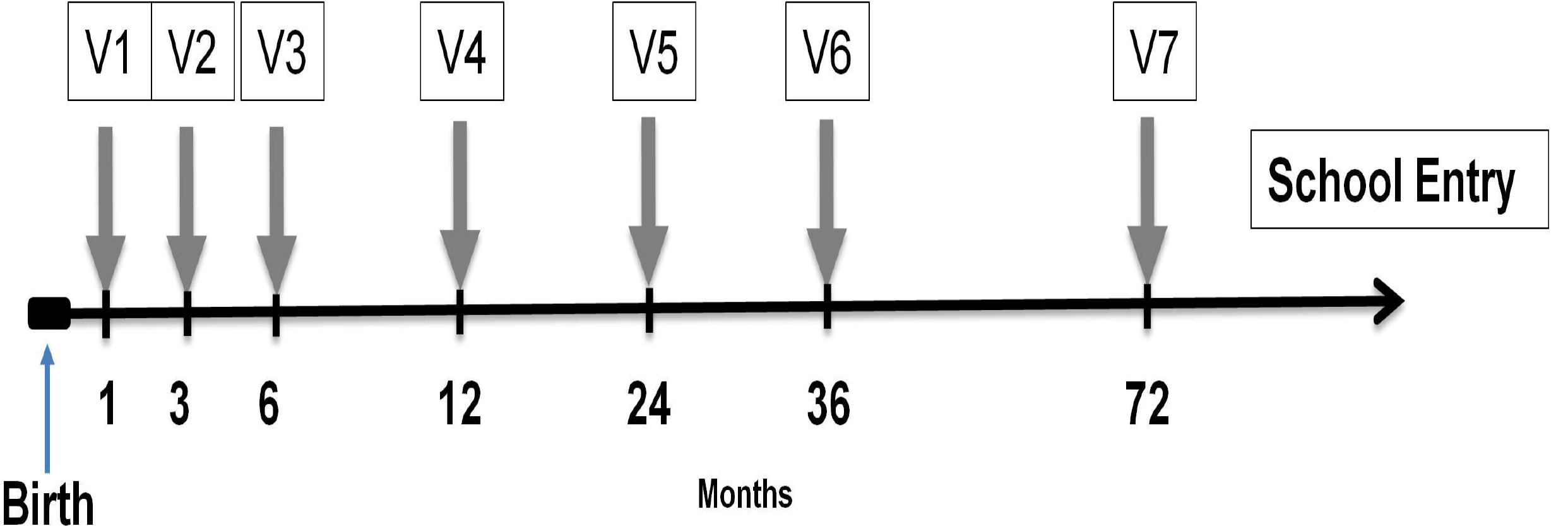
Timeline of data collection in the NASCITA study [11].

Additional visits are guaranteed when needed. The newborn population consists of all infants born during the enrolment period (1 April 2019 – 31 July 2020) and seen by the paediatricians for the well-child visits of the first year of life, if parental consent was given.

Some information was obtained for the NASCITA study in addition to the data routinely collected by the paediatricians during the well-child visits.[11,12]

During the first visit, paediatricians collected sociodemographic data regarding the parents, and information about their health status (e.g. smoking habits, chronic diseases), the pregnancy and the delivery. Moreover, during the well child visits information concerning anthropometric measures of the newborns and feeding habits were collected, and during the 3^rd^ and 4^th^ visits (scheduled at 6 and at 12 months of age, respectively) the paediatricians collected data on infant weaning (timing; traditional versus baby-led), and on the parental perception of appetite and growth of the child. The study was approved by the Fondazione IRCCS Istituto Neurologico “Carlo Besta” ethics committee (Verbale n 59, 6^th^ February 2019,) and informed consent was obtained from the newborns’ parents.

### Outcomes

Infants were classified as underweight (<5^th^ centile), normal (5^th^-85^th^ centile), and overweight (>85^th^ centile) at visit 4 according to the World Health Organization percentiles of body mass index (BMI), estimated on the basis of the gender of the neonate and the age at the moment of the visit.[13]

The likelihood of being overweight/obese at 12 months of age was the primary outcome measure.[14]

### Statistical analysis

The prevalence of overweight/obesity was estimated as the proportion of infants with a BMI >85^th^ centile on all newborns who attended all of the first 4 well-child visits.

For the evaluation of factors associated with overweight/obesity, healthy newborns were selected (i.e. with the exclusion of preterm and low birth weight newborns and of neonates with congenital malformations and/or admitted to intensive care unit). Moreover, the analyses were focused on neonates with appropriate-for-gestational age (AGA) birth weight, estimated using the Italian Neonatal Study (INeS) charts,[15] in order to monitor a cohort with homogeneous characteristics and a similar baseline risk.

Variables associated with an increased risk of overweight/obesity in childhood in previous studies were selected as covariates.[2,3,10]

The covariates considered in the present study were: geographical area of residence (North/Centre/South), residential environment (rural/urban), age of the mother at delivery (<30; 30-34; >34 years), maternal educational level (low: no schooling or primary versus high: secondary school or university), maternal employment status (employed/unemployed), marital status (single yes/no), parity (primiparous/multiparous), nation of birth of the mother (Italy yes/no), presence of maternal chronic diseases, pre-pregnancy BMI, gestational weight gain, type of delivery (spontaneous/caesarean), skin-to-skin contact at partum, gender of the neonate, smoking habits of the mother, and mean number of daily hours spent outdoor by parents and infant (<1; 1-3; >3). Concerning nutrition, we considered: breastfeeding practice (exclusive breastfeeding for at least 6 months, EBF; breastfeeding for at least 12 months, BF), timing of weaning according to the age of the infant in days (<150; 150-179; ≥180), type of weaning (traditional versus baby-led), consumption of commercial baby food (yes/no), parental perception of general infant appetite (poor/normal/big), and parental concern about growth. Traditional weaning involved spoon feeding purees or semi-liquid foods then graduating to more textured foods and some finger foods before joining in with the family diet, while baby-led weaning included different approaches (e.g. “on demand approach/self weaning”, low degree of spoon feeding or puree feeding).[16]

Mothers were grouped according to their pre-pregnancy BMI into three categories, underweight (≤18.5), normal (18.6-24.9), and overweight or obese (≥25.0). To evaluate gestational weight gain, the weight variations recommended by the Institute of Medicine criteria were applied after grouping the mothers according to the pre-pregnancy BMI.[17,18]

Chi-square tests were performed with the aim to evaluate the association between the reported covariates and the outcome measure, and a stepwise logistic regression analysis was performed. Statistical significance was evaluated using a two-tailed P value <0.05. All data management and analyses were performed using SAS software.

## RESULTS

### Prevalence of overweight/obesity at 12 months of age

The NASCITA birth cohort was composed of 5,054 neonates, 3,753 (74.3%) of whom were healthy newborns. For a total of 4,270 subjects (3,420 healthy neonates) data concerning all four well-child visits scheduled during the first year of life were available: 179 (3.5%) children dropped-out, while 605 (12.0%) did not attend one or more well-child visits due to the COVID-19 pandemic situation. The sociodemographic characteristics of the analysed sample (e.g. geographic distribution, maternal age and educational level) were not different from those of the whole cohort.

The prevalence of overweight/obesity was 23.5% (95%CI 22.2-24.8), with no differences when selecting only the healthy newborns (24.2%; 95%CI 22.7-25.7).

### Factors associated with a greater likelihood of being overweight

In all, 2,749 AGA healthy newborns were identified: 1,379 males and 1,370 females. The maternal age at delivery was 33.0 years (SD 5.4), 54.7% of mothers were primiparous, and 12.3% were foreign-born. Almost three fourths (72.4%) of neonates were born via a vaginal delivery. Just over one fourth (26.0%) were exclusively breastfed for at least 6 months, while 45.0% were still being breastfed at 12 months of age. In 46% of the cases, weaning was begun at, or after, six months of age and in 42% it was the baby-led weaning type.

In all, 74 infants (2.7%) resulted underweight at the one-year age visit, 2,024 (73.6%) had a normal BMI, and 651 (23.7%) were overweight or obese.

The geographical area of residence, the age of the mother at delivery, her nationality, educational level and employment status, her pre-pregnancy BMI, the timing and type of weaning, the mean number of daily hours spent outdoors, the parental perception of infant appetite and growth, and the consumption of baby food were associated with different likelihoods of being overweight at 12 months of age at the univariate analyses (Table 1). More specifically, the variables more strongly associated, with the logistic regression analysis, with an increased likelihood of being overweight were an excessive appetite (OR 4.39, 95%CI: 2.73-7.07), living in southern Italy (OR 1.64, 95%CI: 1.35-2.00), the young age of the mother (OR 1.39, 95%CI: 1.05-1.83), and having a foreign-born mother (OR 1.38, 95%CI: 1.05-1.83). The lack of maternal employment and traditional weaning were also associated with a greater likelihood of being overweight (table 2).

**Table 1.**
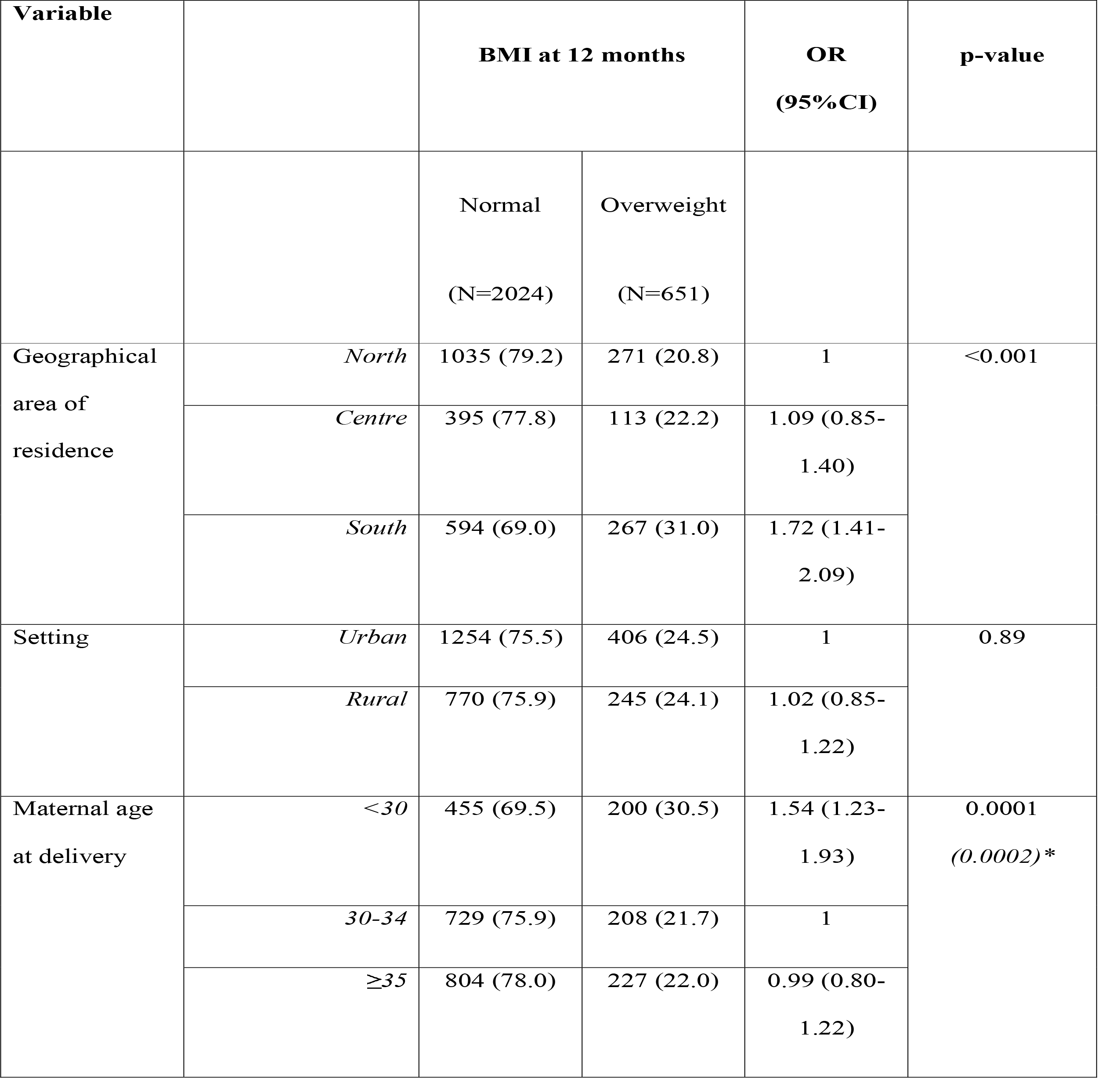
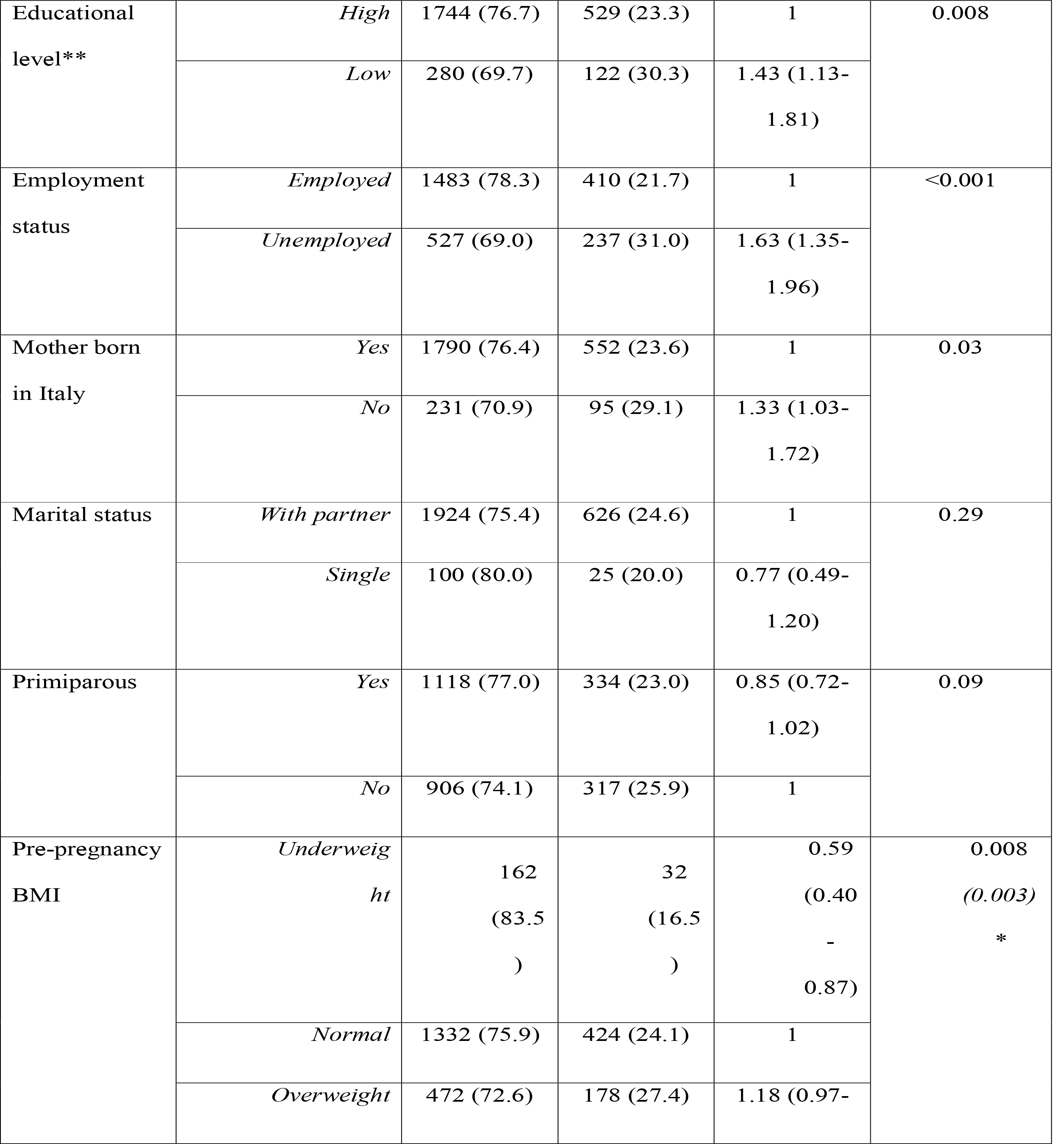
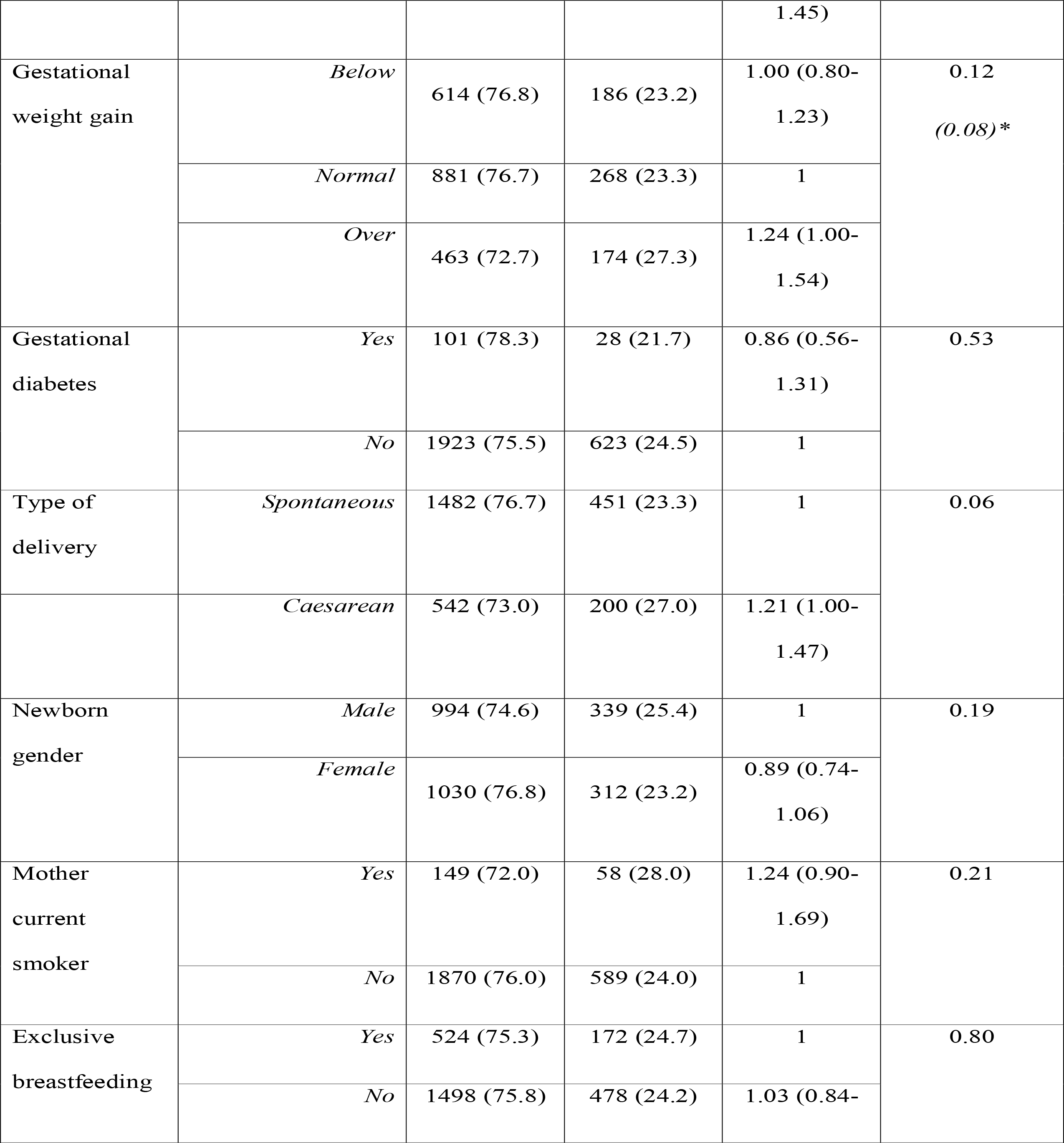
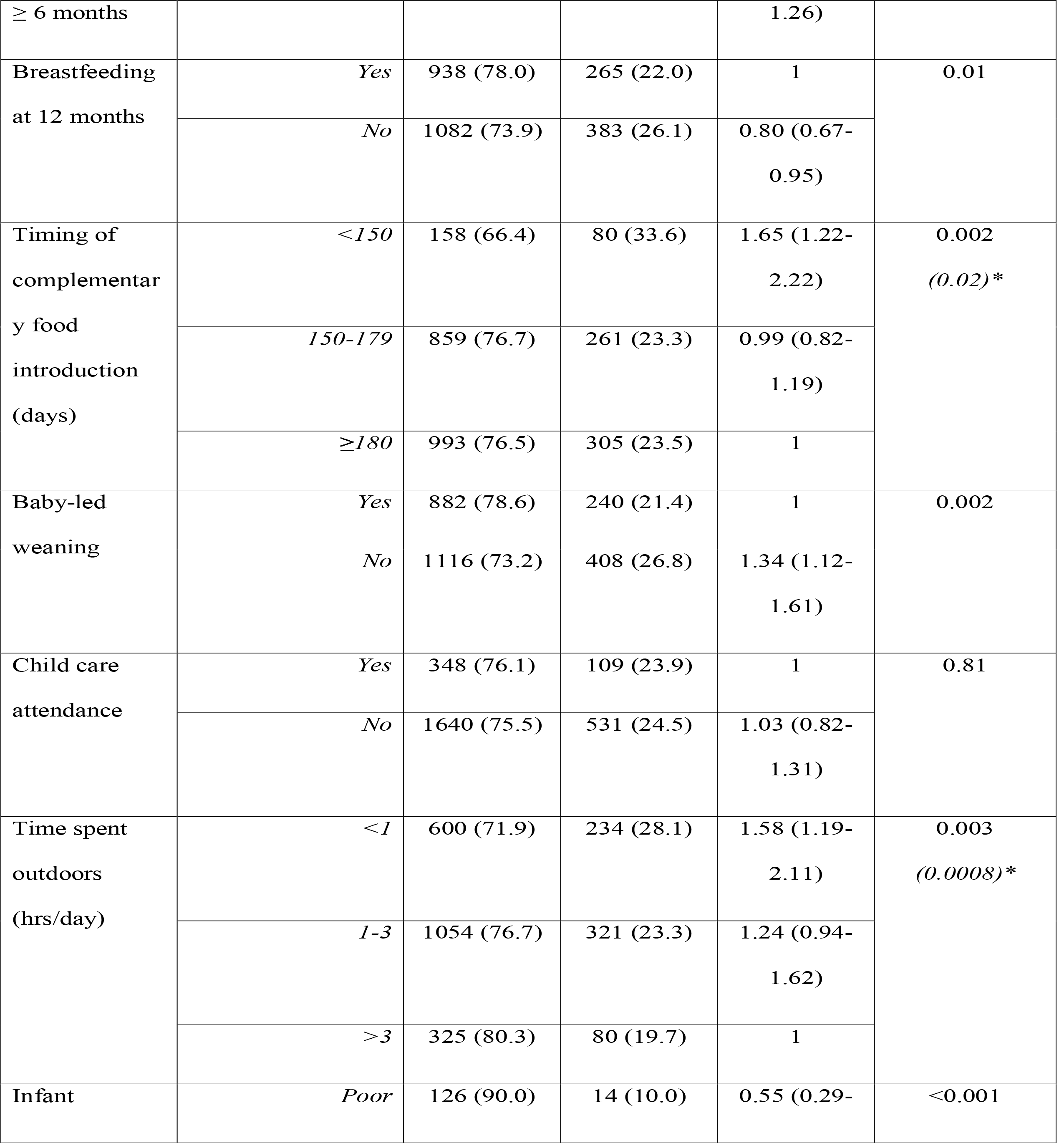
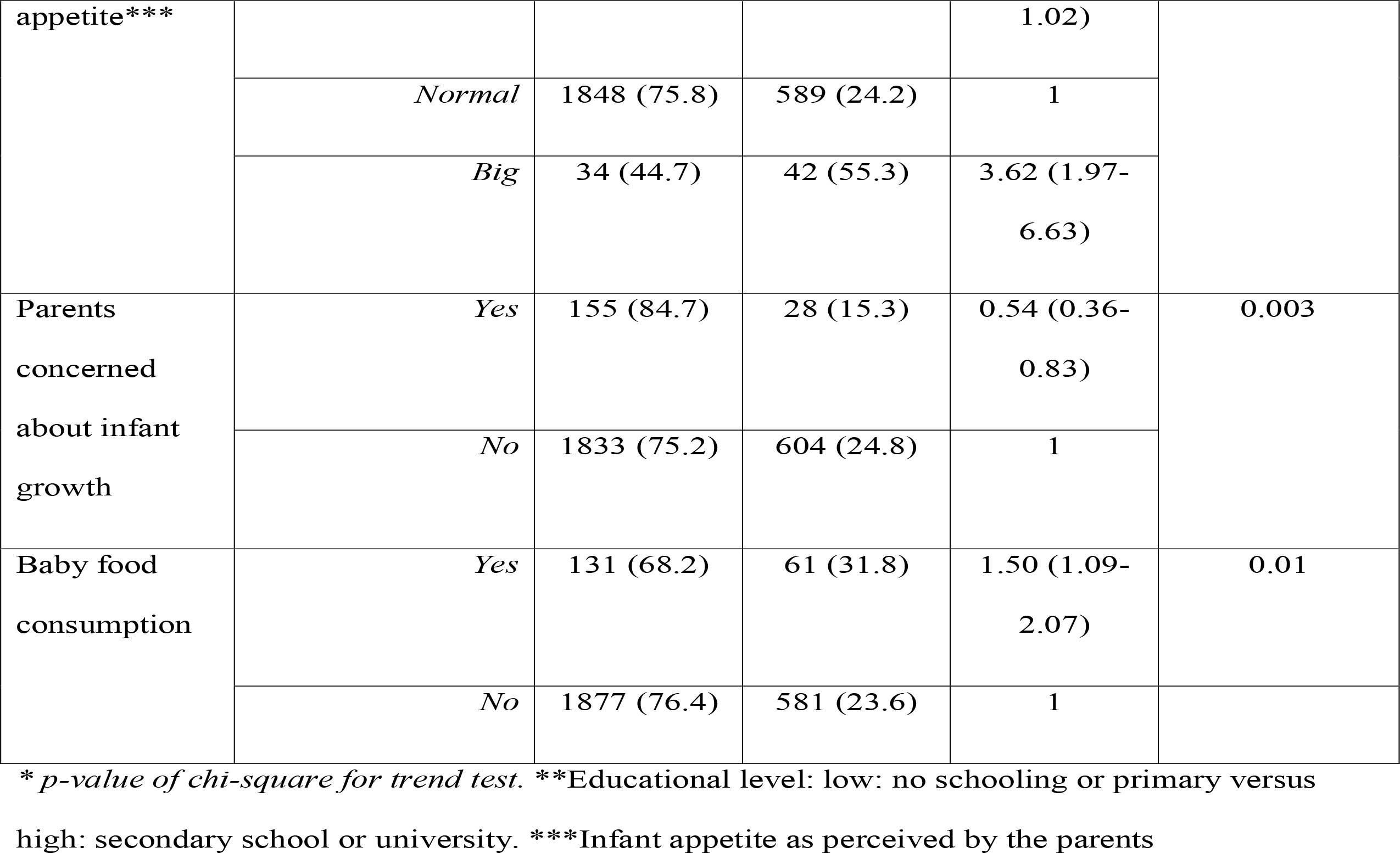
Association between maternal and neonatal characteristics and BMI at 12 months (overweight/obese versus normal).

**Table 2.** Variables associated with overweight/obesity at the stepwise logistic regression analysis.

| **Variable** | **Value** | **Odds Ratio**<br>**(95%CI)\*** | **p-value** |
| --- | --- | --- | --- |
| Geographical area<br>of residence | *North/Centre* | 1 | <0.001 |
|  | *South* | 1.64 (1.35-2.00) |  |
| Maternal age at<br>delivery (yrs) | <30 | 1.39 (1.24 – 1.71) | 0.002 |
|  | 30-34 | 1 | - |
|  | ≥35 | 1.00 (0.80-1.24) | 0.98- |
| Mother born in Italy | *Yes* | 1 | 0.02 |
|  | *No* | 1.38 (1.05-1.83) |  |
| Employment status | *Employed* | 1 | 0.004 |
|  | *Unemployed* | 1.36(1.11-1.67) |  |
| Pre-pregnancy BMI | *Underweight* | 0.58 (0.39-0.87) | 0.008 |
|  | *Normal* | 1 |  |
|  | *Overweight* | 1.10 (0.89-1.36) | 0.41 |
| Baby-led weaning | *Yes* | 1 | 0.009 |
|  | *No* | 1.29 (1.07-1.56) |  |
| Infant appetite* | *Poor* | 0.39 (0.22-0.72) | 0.002 |
|  | *Normal* | 1 | - |
|  | *Big* | 4.39 (2.73-7.07) | <0.001 |

\* General appetite perceived as perceived by the parents

## DISCUSSION

Nearly one out of four infants resulted overweight/obese at 1 year of age according to the WHO growth chart. This finding is the same observed in a cohort of Danish neonates,[19] despite the fact that data reported for school-aged children show a greater prevalence of overweight in the Mediterranean region compared to Northern European countries.[20]

The geographical area of residence emerged as the main factor associated with the risk of overweight, with a 10% difference in prevalence between the North and South of Italy. These data are consistent with those of other studies, which detected a two-fold greater risk of being obese in school-aged children living in the southern regions.[10,21] The fact that geographical differences already emerged at 12 months of age in newborns that had similar characteristics at birth strongly support the relevance of the early months of age in influencing the growth of the child.[22] Italy’s southern regions are characterized by a higher poverty rate, by inequalities in provision of social and educational services, and by a worse status in several health indexes,[23] and this, too, may have an impact on the likelihood of being overweight or obese. The risk of overweight in the South, however, remained significantly higher even after adjusting for several variables. It therefore seems that lower education and socio-economic conditions existing in the South do not fully explain the greatest prevalence of overweight, and that cultural factors, dietary habits, or genetic factors may play a role.

The association between appetite-related traits and overweight in early childhood has already been documented.[24–26]We did not introduce in the NASCITA cohort study a questionnaire for a comprehensive evaluation of the infant eating behaviour (e.g, Baby Eating Behaviour Questionnaire),[27] but we adapted the item concerning the perception of parents about their infant’s appetite. An appetite perceived as “big” by the parents was associated with a 4-fold greater likelihood of being overweight. Llewellyn et al. found that higher birth weight was associated with larger overall appetite,[27] and it is difficult to evaluate if the increased appetite is associated with a greater BMI or vice versa.Our analysis was performed in the AGA subgroup, however, so the influence of birth weight on appetite is reduced.

Young maternal age was one of the other main factors associated with a greater likelihood of overweight. There is no evidence in the literature to support an association between maternal age and offspring overweight,[2] while in our study mothers <30 years old had a greater chance of having overweight infants.

Maternal unemployment was also associated with an increased risk of infant overweight. This is not consistent with other studies, which indicate that full-time employment of the mother increases the chance in children of being overweight.[28–31] It should, however, be considered that most of the studies investigating the association between child weight and maternal employment status involved preschool or school-aged children, while the first year of life is a peculiar situation, since employed mothers are usually on maternity leave for 3 or more months.

Pre-pregnancy overweight and an excessive gestational weight gain have previously been associated with a greater BMI in infancy and childhood.[32–35] This association was not observed in our study, but it is likely that this is due to the choice to include only AGA newborns, while the impact of maternal BMI starts at birth, leading to an increased chance of delivering a large-for-gestational age neonate.[36]

On the contrary, we observed that infants with an underweight mother before pregnancy are less likely to be overweight. This may be partly explained by different dietary habits in underweight mothers.[34]

Despite the fact that evidence is not conclusive, available data suggest that baby-led weaning may be associated with a lower BMI.[37,38] It has been hypothesized that baby-led weaning may improve the infant’s appetite control and lead to higher levels of satiety-responsiveness.[38] This hypothesis is supported also by findings from the NASCITA birth cohort, with a 27% greater likelihood of overweight in children who underwent traditional weaning.

The fact that we observed a lower risk of being overweight when parents were concerned about infant growth seems strange, but it is likely that parents fearing an excess in infant growth may be more sensitive to the quality of the diet.

A few variables resulted as being associated with an increased risk of overweight in the univariate analysis, but not in the logistic regression. In the case of educational level, since unemployed mothers more frequently have a lower educational degree, this could be a confounding variable. No clear association was detected in the available studies between the timing of the introduction of complementary foods and childhood overweight or obesity;[39] in our analysis this association was detected only at the univariate analysis, but not at the multivariate one.

It should be underlined that, with an increase in the average number of hours spent outdoors, there was a decreased risk of overweight, despite the fact that this association was present only in the univariate analysis. It is reasonable to expect that parents who dedicated time for outdoor activities with their infants are also more sensitive to infant nutrition as part of their child’s wellbeing. We did not find an association between breastfeeding (EBF or BF) and overweight, even when testing, as a covariate, the variable ever versus never breastfed (data not shown). Even with conflicting results, breastfeeding is associated with a reduced risk of being overweight.[5–7] Most studies, however, evaluate the risk of overweight/obesity at 2 or 3 years of age, and there are few data concerning infants aged 12 months.[14] We will continue to monitor infant growth in the NASCITA birth cohort to evaluate which variables influence subsequent BMI.

To the best of our knowledge, this is the first study investigating the prevalence of, and the variables related to, overweight at 12 months of age in Italy.

The sample was representative of the national population for distribution by geographical area of residence and environment (rural/urban), and demographic characteristics of the families.[11,12] The anthropometric measures were collected by the paediatricians during the visits and, from this point of view, may be more accurate than when recorded by parents.

Our study has some limitations. First of all, the family paediatricians participated on a voluntary basis and most of them were educated to the best practices for supporting early child development. It is possible that they are not fully representative of Italian paediatricians, and in particular, they may be more sensitive to infant feeding and nutrition.

There were difficulties in data collection due to the COVID-19 pandemic. In particular, well-child visits were in some cases postponed and nearly 10% of infants missed one or more visits due to the parents’ fear of contagion. The characteristics of children included in this analysis were not different from those of the baseline cohort. Moreover, BMI was estimated on the basis of the actual age at the moment of the visit, so a delay of a few weeks had a negligible impact on the data collection.

We were not able to collect information concerning the children’s dietary intake and the quality of their diets. Our definition of baby-led weaning was broad and included different attitudes. We did not, for example, have data concerning the percentage of children receiving spoon feeding or puree feeding.Finally, in the evaluation of factors associated with a greater risk of overweight we choose to include only healthy newborns with an appropriate-for-gestational-age birth weight with the aim to monitor a cohort with the same baseline risk, and our results may apply only to these neonates (the majority). In any case, when considering all healthy newborns independently of their weight-for-gestational-age, the prevalence of overweight/obesity at 12 months was similar to that observed in AGA (24.2% versus 23.7%).

In conclusion, nearly one out of four infants in Italy are overweight/obese. Living in the South and having an young, unemployed mother are the main factors associated with the greatest likelihood of being overweight, while, in our cohort, we did not identify breastfeeding as a potential protective determinant.

It is essential to involve parents in educational interventions, starting from the pregnancy period, aimed at reducing the prevalence of childhood overweight/obesity, taking into account the geographical context (i.e., South of Italy) and the maternal characteristics.[40]

## Data Availability

Data are available from the corresponding author upon reasonable request

## ACKNOWLEDGEMENTS

The authors would like to thank Claudia Pansieri for her collaboration in setting up and running the NASCITA Cohort, Michele Zanetti for the informatics aspects of the setting up and running of the cohort and its website, and Maria Grazia Calati and Daniela Miglio for assistance in management of the cohort and in communication with the paediatricians.

Collaborating author names (NASCITA Working Group): **Scientific Committee:** Antonio Addis, Renata Bortolus, Annalisa Campomori, Sergio Cattani, Luca De Fiore, Federico Marchetti, Cristiana Piloni, Valeria Renzetti, Chiara Segré, Giorgio Tamburlini. **Research area-specific expert collaborators:** Rosario Cavallo, Sergio Conti Nibali, Stefania Manetti, Gherardo Rapisardi, Giacomo Toffol. **Local-level coordinators:** Vicenza Briscioli, Carla Cafaro, Cristoforo Cocchiara, Isodiana Crupi, Patrizia Del Balzo, Laura Dell’edera, Chiara Di Francesco, Alberto Ferrando, Francesca Grassa, Chiara Guidoni, Stefania Manetti, Claudio Mangialavori, Stefano Marinoni, Francesca Marongiu, Fausta Matera, Paolo Moretti, Laura Olimpi, Angela Pasinato, Ilaria Porro, Ippolita Roncoroni, Raffaella Schiro’, Patrizia Seppia, Federica Zanetto. **Participating paediatricians:** Anna Aloisio, Elisabetta Anedda, Giuliana Apuzzo, Giovanna Argo, Anna Armenio, Emanuela Ballerini, Monica Benedetti, Daniela Bertoli, Stefano Bollettini, Chiara Bottalico, Aurora Bottiglieri, Vincenza Briscioli, Antonella Bruno, Laura Brusadin, Mariantonietta Caiazzo, Patrizia Calamita, Miriana Callegari, Rosaria Cambria, Rossella Claudia Cannavò, Maria Cristina Cantù, Domenico Capomolla, Anna Caracciolo, Maria Concetta Carbone, Valeria Carraro, Gaetano Carrassi, Maria Laura Cartiglia, Sara Casagranda, Ornella Castiglione, Rosario Salvatore Cavallo, Teresa Cazzato, Maria Angela Cazzuffi, Melania Maria Giuseppina Cera, Jennifer Chiarolanza, Rosaria China, Nicoletta Cimadamore, Roberto Cionini*, Cristina Ciuffo, Cristoforo Cocchiara, Damiano Colazzo, Sergio Conti Nibali, Anna Maria Costantini, Claudio Cravidi, Marialuisa Criscione, Isodiana Crupi, Rita D’Agostino, Daniela Danieli, Luigi De Carlo, Marina De Sanctis, Giuseppina De Santes, Patrizia Del Balzo, Gian Piero Del Bono, Chiara Di Francesco, Maria Elisabetta Di Pietro, Maria Chiara Dini, Paolo Fiammengo, Micaela Foco, Maria Teresa Fonte, Maria Frigeri, Andrea Galvagno, Matteo Gaudino, Stefania Genoni, Silvia Girotto, Emma Maria Letizia Giugnini, Gianluca Gornati, Marta Gozzi, Francesca Grassa, Chiara Guidoni, Enrica Heritier, Antonella Lavagetto, Raffaele Limauro, Alessandra Magnelli, Maria Gabriella Maiolino, Monica Malventano, Stefania Manetti, Claudio Mangialavori, Silvia Marchi, Natale Maresca, Federico Marolla, Francesca Marongiu, Agata Martinelli, Chiara Martinez, Nicoletta Mascarello, Fausta Matera, Carla Matiotti, Elisabetta Mazzucchi, Donatella Moggia, Paolo Moretti, Manuela Musetti, Paolo Nardini, Alberto Neri, Patrizia Neri, Flavia Nicoloso, Laura Maria Olimpi, Giancarlo Ottonello, Giacinta Padula, Paolo Maria Paganuzzi, Rosanna Palazzi, Alessandra Palmero, Maria Chiara Parisini, Angela Pasinato, Giovannina Pastorelli, Marilena Pavoni, Lucia Peccarisi, Antonella Pellacani, Cristina Perrera, Michela Picciotti, Ivo Picotto, Tiziana Piunti, Ilaria Porro, Francesca Preziosi, Giuseppe Primavera, Miriam Prodi, Maria Letizia Rabbone, Innocenza Rafele, Laura Reali, Franziska Stefanie Rempp, Ada Riundi, Patrizia Rogari, Ippolita Roncoroni, Paolo Rosas, Annarita Russo, Mariagrazia Saccà, Elisabetta Sala, Francesca Sala, Renato Sansone, Francesca Santus, Vittoria Sarno, Alessandra Savino, Raffaella Schirò, Giuseppa Scornavacca*, Giovanni Giuliano Semprini, Maria Francesca Siracusano, Adelisa Spalla, Gloria Sturaro, Giacomo Toffol, Maria Grazia Toma, Ettore Tomagra, Maria Tortorella, Fausta Trentadue, Marina Trevisan, Silvia Tulisso, Roberta Usella, Anna Valente, Michele Valente, Mariangela Valera, Edda Vernile, Valeria Vicario, Lucia Vignutelli, Paolo Vinci, Lucia Vizziello, Rosette Zand, Marco Zanette, Federica Zanetto, Graziano Zucchi, Maria Luisa Zuccolo. Roberto Cionini and Giuseppa Scornavacca unfortunately passed away.

## FUNDING

This work was supported by resources from the Laboratory for Mother and Child Health and by an economic contribution by the *Associazione Amici del Mario Negri*.

The *Associazione Amici del Mario Negri* had no role in the design and conduct of the study.

## DATA AVAILABILITY STATEMENT

Data are available from the corresponding author upon reasonable request.

## ETHICS STATEMENTS

### Patient consent for publication

Not applicable.

### Ethics approval

The study was approved by the Fondazione IRCCS Istituto Neurologico “Carlo Besta” ethics committee (Verbale n 59, 6th February 2019) and informed consent was obtained from the newborns’ parents.

